# Sleep and Its Cardiovascular and Cognitive Influences in the Academic Environment

**DOI:** 10.64898/2026.01.08.26343729

**Authors:** Laura Cecilia Fernandes Silva, Andreza Vieira Ramos, Fernanda Follis Tasso, Rafael Teixeira Hurtado, Ândrea Gabrielle Batistela da Silva, Victoria Giulia Ramos Silveira, Leonardo Trindade Rodrigues, Moacir Fernandes de Godoy

## Abstract

**Introduction:** Sleep is a vital physiological process essential for memory consolidation, cognitive performance, and the regulation of cardiovascular functions. Students in health sciences programs constitute a population particularly susceptible to sleep disturbances due to heavy academic workloads, irregular schedules, and psychosocial stressors.

**Objective:** To investigate the relationship between sleep patterns, academic performance, and cardiovascular risk factors among health sciences students.

**Methods:** A cross-sectional study was conducted involving 349 students enrolled in Medicine, Psychology, and Nursing programs. Participants completed a sociodemographic questionnaire, the Pittsburgh Sleep Quality Index (PSQI), and the Epworth Sleepiness Scale (ESS). Anthropometric data, including weight, height, waist circumference, and blood pressure were measured. Statistical analyses comprised descriptive and inferential methods.

**Results:** A high prevalence of poor sleep quality and excessive daytime sleepiness was identified. Sleep quality was significantly associated with academic performance (p = 0.003). Male sex, smoking, and higher body mass index (BMI) were also correlated with poorer sleep patterns.

**Conclusion:** Poor sleep quality adversely affects academic performance and is associated with cardiovascular risk factors. The implementation of institutional strategies aimed at promoting sleep hygiene and providing psychosocial support is recommended to mitigate these impacts.

## Introduction

Sleep plays an essential role in maintaining physical and mental health, being a key determinant of processes such as memory consolidation, emotional regulation, cognitive performance, and cardiovascular stability (1). In the academic setting, however, inadequate sleep habits are common, often driven by prolonged study periods, demanding deadlines, and high levels of stress (2,3). This context favors sleep deprivation and fragmentation, with potential negative consequences for both cognition and the cardiovascular system (4–6).

Evidence indicates that poor sleep quality impairs attention, learning, and academic achievement. Furthermore, sleep disorders are associated with alterations in autonomic regulation, elevated blood pressure, metabolic dysfunction, and an increased risk of cardiovascular diseases (7). Despite these findings, the impact of sleep on university students—who are exposed to multiple psychosocial risk factors—remains frequently underestimated (8).

Far from representing a passive state of rest, sleep is an active and highly regulated process mediated by complex neurochemical mechanisms (9,10). During non–rapid eye movement (NREM) and rapid eye movement (REM) sleep stages, critical physiological functions necessary for somatic and mental homeostasis take place (11). Over recent decades, research has consistently demonstrated a link between sleep deprivation and cognitive impairment, metabolic disturbances, and cardiovascular dysfunction. Among the most relevant sleep disorders, obstructive sleep apnea stands out, as it is associated with hypertension, obesity, insulin resistance, and an increased risk of cardiac events.

This scenario is of particular relevance in the university context. Students face challenges such as intense academic demands, excessive use of digital technologies, and irregular daily routines, all of which exacerbate sleep deprivation (12). Such conditions directly affect essential cognitive functions, including memory, attention, and reasoning, and also pose a health risk to the cardiovascular system of this young population (13).

The selection of health-related degree programs (Medicine, Nursing, and Psychology) is justified by their high academic demands, extended workloads, and early exposure to stressful situations and human suffering, factors that contribute to dysregulated sleep patterns and increased vulnerability to their physiological and cognitive consequences (14,15).

Although international studies have addressed the impact of sleep on university students’ health, there is a scarcity of national data integrating variables such as sleep quality, cardiovascular risk, and academic performance, particularly in public higher education institutions located in inland regions of Brazil.

In view of the relevance of the interactions between sleep, cognition, and cardiovascular health—especially within a highly demanding academic environment—this study aims to evaluate sleep quality and duration among students of Medicine, Nursing, and Psychology at the Faculdade de Medicina de São José do Rio Preto (FAMERP), and to correlate these data with cardiovascular risk factors and academic performance.

To this end, an observational study was conducted using validated instruments to measure sleep quality and duration, identify cardiovascular risk factors, and analyze participants’ academic performance. It is hypothesized that students with poorer sleep quality will present a higher prevalence of cardiovascular risk factors and lower academic performance. This research seeks to provide an updated overview of the impact of sleep in this population and to contribute to awareness and health promotion strategies based on scientific evidence.

## Methods

This is a cross-sectional, observational, individual-level study approved by the Research Ethics Committee of the Faculdade de Medicina de São José do Rio Preto (FAMERP), under CAAE number 79790024.3.0000.5415, in compliance with the ethical principles established by Resolution No. 466/2012 of the Brazilian National Health Council.

Data collection was carried out between September and November 2024 at FAMERP, a public higher education institution located in the interior of the state of São Paulo, Brazil. The sample included students regularly enrolled in the Medicine, Nursing, and Psychology programs. Data tabulation and statistical analyses were conducted between December 2024 and February 2025, followed by discussions in March and final manuscript preparation in June.

Data was collected in person. The inclusion criteria were: being enrolled in one of the three programs and being 18 years of age or older. There were no exclusion criteria. Each participant received a printed questionnaire along with the Informed Consent Form, containing items related to cardiovascular risk factors, personal history of arterial hypertension, diabetes mellitus, and hypercholesterolemia, as well as lifestyle habits, sleep quality, and academic performance. Blood pressure and abdominal circumference were also measured using the same blood pressure device and measuring tape for all participants.

Family history included diagnoses of arterial hypertension, diabetes mellitus, and hypercholesterolemia, which are relevant familial cardiovascular risk factors (16). For statistical analysis, data were grouped according to the isolated or combined presence of these conditions. Personal medical history was also evaluated for the same conditions. Sociocultural practices, including alcohol consumption, smoking, and physical activity, were analyzed and dichotomized as absent or present at levels above 150 minutes per week, in accordance with World Health Organization (WHO) guidelines (17).

Body mass index (BMI) was calculated using self-reported weight and height obtained from the questionnaire. For statistical analysis, six BMI categories were considered: underweight (BMI < 18.5 kg/m²), normal weight (BMI 18.5–24.9 kg/m²), overweight (BMI 25.0–29.9 kg/m²), obesity class I (BMI 30.0–34.9 kg/m²), obesity class II (BMI 35.0–39.9 kg/m²), and obesity class III (BMI ≥ 40.0 kg/m²).

Blood pressure was measured in mmHg in all participants; however, no standardized time of day was established for the measurements, and no formal training for measurement standardization was conducted. Different sphygmomanometers were used, which had not undergone calibration procedures; however, all devices were certified by the Brazilian National Institute of Metrology (INMETRO). Abdominal circumference was measured in centimeters using the same model of measuring tape, although no formal standardization training was performed.

To assess academic performance, participants reported their annual grade point averages. Verification of these data was not possible, as access to the institutional academic system was not available. For statistical purposes, participants were stratified into two groups: grade point averages equal to or below 6.5 (considering that the institutional passing average is 6.5) and grade point averages above 6.5. Participants were also asked whether they had ever been required to take a remedial/second-chance examination due to insufficient grades and how many times this had occurred. For statistical analysis, this variable was dichotomized into those who had and those who had not required such examinations.

Sleep quality was assessed using two validated instruments: the Pittsburgh Sleep Quality Index (18) and the Epworth Sleepiness Scale (19). The former includes items regarding usual bedtime and wake-up time, sleep latency, total sleep duration, and external factors that may interfere with sleep quality, such as temperature, dyspnea, nightmares, and pain. The latter focuses on the assessment of daytime sleepiness, with questions regarding the likelihood of dozing off in different situations, such as watching television, reading, and talking. Both scales were administered to all participants and were used in the statistical analyses to compare sleep parameters with cognitive performance and cardiovascular risk factors.

## Results

After data collection, the information was organized in Microsoft Excel spreadsheets and subsequently analyzed using the Statistical Package for the Social Sciences (SPSS, version 24.0), GraphPad InStat (version 3.10, 2009), and Prism (version 6.07, 2015). Descriptive statistical analysis included measures of central tendency (mean, median) and dispersion (standard deviation), as well as absolute and relative frequency distributions for categorical variables. For inferential analysis, the normality of quantitative variables was assessed using the Kolmogorov–Smirnov test. According to data distribution, appropriate parametric or nonparametric tests were applied for group comparisons. Proportions were compared using the chi-square test and Fisher’s exact test when necessary. Correlations between quantitative variables were evaluated using Pearson’s or Spearman’s correlation coefficients, depending on data characteristics. A significance level of 5% (p ≤ 0.05) was adopted for all analyses.

The sample consisted of 349 individuals, with a mean BMI of 24.09 (SD = 3.91) and a median of 23.55, ranging from 16.00 to 41.52. Regarding waist circumference, 76.3% were classified as low cardiovascular risk (152 women and 115 men) and 23.7% as high risk, with a higher prevalence among women (68.7%) than men (31.3%). Most participants presented blood pressure within normal ranges (75.1%), while 87 individuals (24.9%) had elevated blood pressure, slightly more prevalent among women (54%) than men (46%).

With respect to physical activity, 87.1% of the sample reported engaging in physical activity, whereas 12.9% reported no regular activity, with inactivity being more frequent among women (80%) compared with men (20%). Regarding academic program, most participants were enrolled in Medicine (55.4%), followed by Nursing (29.7%) and Psychology (14.9%), with a higher proportion of men in the Medicine program (58.8%) and a predominance of women in Nursing (87.5%) and Psychology (73%). Alcohol consumption was reported by 61.1% of participants, being more prevalent among women (57%) than men (43%). Most participants were non-smokers (90.9%), while 9.1% reported smoking, with smoking being more common among men (56.25%) than women (43.75%). These findings characterize the sociodemographic and health profile of the sample, showing a predominance of females and generally favorable health indicators, particularly in relation to physical activity and low smoking rates.

When comparing sleep quality and academic performance, a significant association was observed (p = 0.003), suggesting that individuals with poor sleep quality tend to present poorer academic performance. Of the 349 participants, 74 had poor sleep quality and 275 had satisfactory sleep quality. Among those with poor sleep, only one presented satisfactory academic performance, whereas 32 had unsatisfactory performance. Among those with good sleep quality, 73 had satisfactory performance and 243 had unsatisfactory performance.

In contrast, the analysis of daytime sleepiness and academic performance (p = 0.6625) indicated that the presence of daytime sleepiness was not associated with worse academic performance. Of the 349 participants, 182 reported daytime sleepiness and 167 did not. Among those with excessive sleepiness, 16 had satisfactory academic performance and 166 had unsatisfactory performance. Among those without sleepiness, 17 had satisfactory performance and 150 had unsatisfactory performance.

Table 1 shows the relationship between sleep quality, daytime sleepiness, and several variables through p-values. No statistically significant associations were observed between sleep quality and any of the analyzed variables, including BMI (p = 0.172), waist circumference (p = 0.1615), blood pressure (p = 0.7285), physical activity (p = 0.5715), academic program (p = 0.6062), and sex (p = 0.9564). Conversely, daytime sleepiness showed a statistically significant association with sex (p = 0.0006), indicating that sleepiness varied according to participants’ sex. The other variables—BMI (p = 0.1351), waist circumference (p = 0.4789), blood pressure (p = 0.2107), physical activity (p = 0.872), and academic program (p = 0.1902)—were not significantly associated with daytime sleepiness.

**Table 1.** General characteristics of the sample. Source: study data.

| Variable | Result |
| --- | --- |
| Number of participants | 349 |
| Sex | 65% female |
| Mean age | 22,4 ± 2,3 years |
| Poor sleep quality (PSQI > 5) | 53% |
| Excessive daytime sleepiness (ESS ≥ 10) | 37% |

Table 2 presents the relationship between daytime sleepiness and variables of interest, including alcohol consumption, smoking, physical activity, BMI, blood pressure, and waist circumference. Alcohol consumption showed an odds ratio (OR) of 0.9474 (95% CI: 0.6073–1.47919; p = 0.8132), indicating no statistically significant association. Smoking showed an OR of 1.07978 (95% CI: 0.51366–2.24423; p = 0.8084), also without statistical significance. The absence of physical activity had an OR of 0.66596 (95% CI: 0.34801–1.19933; p = 0.2034), suggesting a non-significant trend. BMI showed an OR of 0.954168; however, the reported confidence interval was inconsistent, and the p-value (0.1395) suggested only potential clinical relevance. Blood pressure (OR = 1.1104; 95% CI: 0.58384–2.13108; p = 0.7371) and waist circumference (OR = 1.1104; 95% CI: 0.58384–2.13108; p = 0.7371) showed no statistically significant associations.

**Table 2.** Association between daytime sleepiness and clinical and behavioral variables (multivariable model). Source: study data.

| Variable | OR | 95% CI | p-value |
| --- | --- | --- | --- |
| Alcohol consumption<br>(yes vs. no) | 0,9474 | 0,6073–1,47919 | 0,8132 |
| Smoking (yes vs. no) | 1,07978 | 0,51366–2,24423 | 0,8084 |
| Physical activity<br>(absence) | 0,66596 | 0,34801–1,19933 | 0,2034 |
| BMI (continuous) | 0,954168 | 0,640228–1,47675 | 0,1395 |
| High blood pressure | 1,1104 | 0,58384–2,13108 | 0,7371 |
| Waist<br>circumference<br>(high risk) | 1,1104 | 0,58384–2,13108 | 0,7371 |

Table 3 shows the multivariate relationship between daytime sleepiness and selected variables (physical activity, BMI, blood pressure, and waist circumference). Model fit tests indicated statistical significance (deviance goodness-of-fit chi-square = 38.74484, df = 27, p < 0.0001; likelihood ratio chi-square = 57.5409, df = 2, p = 0.0053). The absence of physical activity showed an OR of 0.65347 (95% CI: 0.34552–1.24362; p = 0.1837), suggesting a non-significant clinical trend. BMI showed an OR of 0.94732 (95% CI: 0.59678–1.00176; p = 0.0551), indicating a borderline association with reduced risk of daytime sleepiness as BMI increased.

**Table 3.** Reduced model for daytime sleepiness (physical activity and BMI). Source: study data.

| Variable | OR | 95% CI | p-value |
| --- | --- | --- | --- |
| Physical activity<br>(absence) | 0,65347 | 0,34552–1,24362 | 0,1837 |
| BMI (continuous) | 0,94732 | 0,59678–1,00176 | 0,0551 |

Table 4 presents the multivariate analysis of sleep quality and variables of interest. Model fit tests indicated no statistically significant model fit (deviance goodness-of-fit chi-square = 33.50201, df = 31, p = 0.3455; likelihood ratio chi-square = 7.38071, df = 6, p = 0.2889). Alcohol consumption showed an OR of 1.2865 (95% CI: 0.744015–2.11973; p = 0.3679). Smoking showed an OR of 0.37765 with an inconsistent confidence interval and p = 0.0848, suggesting only a clinical trend. The absence of physical activity showed an OR of 1.48792 (95% CI: 0.661074–3.10195; p = 0.3107). BMI (OR = 0.96726; 95% CI: 0.88724–1.05807; p = 0.4695) and blood pressure (OR = 1.17544; 95% CI: 0.558165–2.41365; p = 0.671) were not significantly associated. Waist circumference showed an OR of 1.84972 (95% CI: 0.73197–4.71797; p = 0.1579), suggesting potential clinical relevance.

**Table 4.** Association between sleep quality and clinical and behavioral variables (multivariable model). Source: study data.

| Variable | OR | 95% CI | p-value |
| --- | --- | --- | --- |
| Alcohol consumption<br>(yes vs. no) | 1,2865 | 0,744015–2,11973 | 0,3679 |
| Smoking (yes vs. no) | 0,37765 | 0,09834–1,72371 | 0,0848 |
| Physical activity<br>(absence) | 1,48792 | 0,661074–3,10195 | 0,3107 |
| BMI (continuous) | 0,96726 | 0,88724–1,05807 | 0,4695 |
| High blood pressure | 1,17544 | 0,558165–2,41365 | 0,6710 |
| Waist<br>circumference<br>(high risk) | 1,84972 | 0,73197–4,71797 | 0,1579 |

Finally, Table 5 shows that no statistically significant associations were observed between sleep quality and the cardiovascular risk factors evaluated (smoking and waist circumference). Smoking presented an OR of 0.375 (95% CI: 0.11–1.27; p = 0.116), and waist circumference presented an OR of 1.53 (95% CI: 0.79–2.97; p = 0.2025). Model goodness-of-fit tests (p = 0.7654 and p = 0.0754) indicated satisfactory model fit; however, the included variables did not significantly explain variability in sleep quality in the analyzed sample.

**Table 5.** Reduced model for sleep quality (smoking and waist circumference). Source: study data.

| Variable | OR | 95% CI | p-value |
| --- | --- | --- | --- |
| Smoking (yes vs. no) | 0,375 | 0,11–1,27 | 0,116 |
| Waist circumference<br>(high risk) | 1,53 | 0,79–2,97 | 0,2025 |

## Discussion

Taken together, the findings suggest that daytime sleepiness and poor sleep quality among university students result from multifactorial determinants, involving both individual characteristics and academic and social pressures. Institutional policies that promote sleep hygiene, flexible timetables, and psychopedagogical support may help mitigate these negative impacts and improve academic performance.

Sex-related differences also deserve attention. A higher prevalence of daytime sleepiness was observed among women, which is consistent with studies reporting poorer subjective perception of sleep quality in females, possibly influenced by hormonal variations, social overload, and sleep fragmentation (20,21). Conversely, objective assessments indicate a higher frequency of obstructive sleep apnea and nocturnal awakenings in men, reinforcing that both physiological and behavioral factors modulate sleep differently according to sex.

Another relevant aspect is smoking, which in this study showed only a clinical trend toward association with poorer sleep quality, without reaching statistical significance. Nevertheless, polysomnographic studies have demonstrated that smokers present longer sleep latency, shorter total sleep time, and more frequent nocturnal awakenings, suggesting a biologically plausible effect of nicotine on sleep fragmentation (22,23).

The result related to body mass index (BMI) showed an inverse trend with daytime sleepiness, diverging from much of the literature, which identifies excess body weight as a risk factor for sleep-disordered breathing and hypersomnolence (24,25). A possible explanation is that the sample was predominantly composed of young, normal-weight, and physically active individuals, in whom psychosocial factors such as academic stress and irregular sleep schedules may exert a greater influence than metabolic alterations. This finding reinforces the need to investigate interactions between body composition, lifestyle habits, and sleep patterns in university populations.

Sleep quality has been increasingly recognized as a central determinant of physical, emotional, and academic health in university students, particularly in health-related programs. In the present study, we identified a statistically significant association between poor sleep quality and poorer academic performance, a finding consistent with systematic reviews demonstrating that sleep deprivation or fragmentation impairs attention, memory, and learning in medical and nursing students (26,27).

## Conclusion

In light of these findings, it is recommended that higher education institutions incorporate sleep health promotion actions into their curricula, including guidance on sleep hygiene, stress management, and monitoring of risk factors. Preventive interventions may not only improve students’ quality of life and cardiovascular health but also support their academic and professional training. This study demonstrated a high prevalence of poor sleep quality and its association with worse academic performance among health sciences students. Although excessive daytime sleepiness was not significantly correlated with academic grades, trends were observed suggesting the influence of factors such as sex, smoking, and body composition. These results reinforce the complexity of the relationship between sleep and cognition, in which physiological, psychological, and social aspects interact.

## Ethical Aspects

Name of the Research Ethics Committee that approved the study: Faculdade de Medicina de São José do Rio Preto – FAMERP, São Paulo, Brazil

Certificate of Ethical Review Submission (CAAE): 79790024.3.0000.5415

Informed consent: Obtained from all participants before the start of the study.

### List of abbreviations

PSQI: Pittsburgh Sleep Quality Index
ESS: Epworth Sleepiness Scale
BMI: Body Mass Index
HAS: Systemic Arterial Hypertension
DM: Diabetes Mellitus
HC: Hypercholesterolemia

## Data Availability

All data produced in the present work are contained in the manuscript.

## References

1. Suardiaz-Muro M, Morante-Ruiz M, Ortega-Moreno M, Ruiz MA, Martín-Plasencia P, Vela-Bueno A. Sleep and academic performance in university students: a systematic review. Rev Neurol. 2020;71(2):43-53. doi:10.33588/rn.7102.2020015

2. Wu TT, Zou YL, Xu KD, Jiang XR, Zhou MM, Zhang SB, et al. Insomnia and multiple health outcomes: umbrella review of meta-analyses of prospective cohort studies. Public Health. 2023;215:66-74. doi:10.1016/j.puhe.2022.11.021

3. Goldsborough E 3rd, Osuji N, Blaha MJ. Assessment of cardiovascular disease risk: a 2022 update. Endocrinol Metab Clin North Am. 2022 Sep;51(3):483–509. doi: 10.1016/j.ecl.2022.02.005. PMID: 35963625

4. Camargo EM, Añez CRR. Diretrizes da OMS para atividade física e comportamento sedentário, num piscar de olhos. Genebra: Organização Mundial da Saúde; 2020. Disponível em: https://apps.who.int/iris/handle/10665/337001

5. Zitser J, Allen IE, Falgàs N, Le MM, Neylan TC, Kramer JH, et al. Pittsburgh Sleep Quality Index (PSQI) responses are modulated by total sleep time and wake after sleep onset in healthy older adults. PLoS One. 2022 Jun 24;17(6):e0270095. doi: 10.1371/journal.pone.0270095. PMID: 35749529

6. Lok R, Zeitzer JM. Physiological correlates of the Epworth Sleepiness Scale reveal different dimensions of daytime sleepiness. Sleep Adv. 2021 May 29;2(1):zpab008. doi: 10.1093/sleepadvances/zpab008. PMID: 34250482

7. Miletínová E, Bušková J. Functions of sleep. Physiol Res. 2021 Apr 30;70(2):177–82. doi: 10.33549/physiolres.934470. PMID: 33676389

8. Gardani M, Bradford DRR, Russell K, Allan S, Beattie L, Ellis JG, et al. A systematic review and meta-analysis of poor sleep, insomnia symptoms and stress in undergraduate students. Sleep Med Rev. 2022 Feb;61:101565. doi: 10.1016/j.smrv.2021.101565. PMID: 34922108

9. Sarode R, Nikam PP. The impact of sleep disorders on cardiovascular health: mechanisms and interventions. Cureus. 2023 Nov 30;15(11):e49703. doi: 10.7759/cureus.49703. PMID: 38161933

10. Silva RP, Santos AM, Almeida LM. Fatores associados à qualidade do sono de estudantes universitários. Cienc Saude Coletiva. 2023 Apr;28(4):1413–22. doi: 10.1590/1413-81232023284.14132022

11. Perotta B, Arantes-Costa FM, Enns SC, Figueiro-Filho EA, Paro H, Santos IS, et al. Sleepiness, sleep deprivation, quality of life, mental symptoms and perception of academic environment in medical students. BMC Med Educ. 2021 Feb 17;21(1):111. doi: 10.1186/s12909-021-02544-8. PMID: 33596885

12. Holst SC, Landolt HP. Sleep-wake neurochemistry. Sleep Med Clin. 2022;17(2):151–60. doi: 10.1016/j.jsmc.2022.03.002

13. Khan MS, Aouad R. The effects of insomnia and sleep loss on cardiovascular disease. Sleep Med Clin. 2022 Jun;17(2):193–203. doi: 10.1016/j.jsmc.2022.02.008. PMID: 35659073

14. Miller MA, Howarth NE. Sleep and cardiovascular disease. Emerg Top Life Sci. 2023 Dec 22;7(5):457–66. doi: 10.1042/ETLS20230111. PMID: 38084859

15. Sánchez-de-la-Torre M, Barbé F. Sleep disorders and cardiovascular disease. Med Clin (Barc). 2022 Jan 21;158(2):73–5. doi: 10.1016/j.medcli.2021.09.001. PMID: 34627601

16. Walker MP. The role of sleep in brain function and systemic physiology. Annu Rev Neurosci. 2021 Jul;44:345–68. doi: 10.1146/annurev-neuro-091420-112111

17. Becker SP, Dvorsky MR. Sleep and mental health in college students: mechanisms and opportunities for intervention. Curr Opin Psychol. 2023 Dec;48:101456. doi: 10.1016/j.copsyc.2023.101456

18. Chellappa SL, Aeschbach D. Sleep and cognitive performance in young adults: implications for cardiovascular health. J Sleep Res. 2022 Aug;31(4):e13567. doi: 10.1111/jsr.13567

19. Almeida LS, Cruz JF. Sleep patterns and stress in health sciences students: a cross-sectional study in medical, nursing, and psychology undergraduates. BMC Med Educ. 2023 Apr;23(1):245. doi: 10.1186/s12909-023-04123-8

20. Dyrbye LN, Shanafelt TD. Sleep deprivation and burnout among medical students: a longitudinal study. Acad Med. 2022 May;97(5):698–706. doi: 10.1097/ACM.0000000000004612

21. Cohrs S, Rodenbeck A, Riemann D, et al. Impaired sleep quality and sleep duration in smokers-results from the German Multicenter Study on Nicotine Dependence. Addict Biol. 2014;19(3):486–496. doi:10.1111/j.1369-1600.2012.00487.x

22. Jaehne A, Unbehaun T, Feige B, Lutz UC, Batra A, Riemann D. How smoking affects sleep: a polysomnographical analysis. Sleep Med. 2012;13(10):1286–1292. doi:10.1016/j.sleep.2012.06.026

23. MOREIRA, L. P., et al. Comparação da qualidade do sono entre homens e mulheres ativos fisicamente. Revista de Estudos em Saúde Coletiva, v. 2, p. 38–49, 2013

24. SILVA, A. et al. Diferenças de gênero e idade nos resultados da polissonografia e nas queixas de sono de pacientes encaminhados a um laboratório do sono. Jornal Brasileiro de Pneumologia, 2008

25. Lima PF, Medeiros AL, Araujo JF. Sleep-wake pattern of medical students: early versus late class starting time. Braz J Med Biol Res. 2002;35(11):1373–7

26. Buysse DJ, Reynolds CF, Monk TH, Berman SR, Kupfer DJ. The Pittsburgh Sleep Quality Index: a new instrument for psychiatric practice and research. Psychiatry Res. 1989;28(2):193–213

27. Johns MW. A new method for measuring daytime sleepiness: the Epworth Sleepiness Scale. Sleep. 1991;14(6):540–5

